# Effect of Time to Thrombolysis on Clinical Outcomes in Patients with Acute Ischemic Stroke Treated with Tenecteplase Compared to Alteplase: Analysis from the AcT Randomized Controlled Trial

**DOI:** 10.1101/2023.05.17.23290141

**Authors:** Nishita Singh, Mohammed Almekhlafi, Fouzi Bala, Ayoola Ademola, Shelagh B. Coutts, Yan Deschaintre, Houman Khosravani, Brian Buck, Ramana Appireddy, Francois Moreau, Gord Gubitz, Aleksander Tkach, Luciana Catanese, Dar Dowlatshahi, George Medvedev, Jennifer Mandzia, Aleksandra Pikula, Jai Jai Shankar, Esseeddeegg Ghrooda, Alexandre Y. Poppe, Heather Williams, Thalia S. Field, Alejandro Manosalva, Muzaffar Siddiqui, Atif Zafar, Oje Imoukhoude, Gary Hunter, Michel Shamy, Andrew M. Demchuk, Brian Claggett, Michael D. Hill, Tolulope T. Sajobi, Richard H. Swartz, Bijoy K. Menon

## Abstract

**Background:** The Alteplase compared to Tenecteplase (AcT) randomized controlled trial (RCT) showed that tenecteplase is non-inferior to alteplase in treating acute ischemic stroke within 4.5 hours of symptom onset. The effect of time to treatment on clinical outcomes with alteplase is well known, however the nature of this relationship is yet to be described with tenecteplase. We assessed whether the association of time to thrombolysis treatment with clinical outcomes in patients with acute ischemic stroke differs by whether they receive intravenous tenecteplase versus alteplase.

**Methods:** Patients included were from AcT, a pragmatic, registry linked, phase 3 RCT comparing intravenous tenecteplase to alteplase in patients with acute ischemic stroke. Eligible patients were >18 years old, with disabling neurological deficits, presenting within 4·5 hours of symptom onset, and eligible for thrombolysis. Primary outcome was modified Rankin scale(mRS) 0-1 at 90 days. Safety outcomes included 24-hour symptomatic intracerebral hemorrhage (sICH) and 90-day mortality rates. Mixed effects logistic regression was used to assess a)the association of stroke symptom onset to needle time (ONT), b)door (hospital arrival) to needle time(DNT) with outcomes and c)if these associations were modified by type of thrombolytic administered (tenecteplase vs. alteplase), after adjusting for age, sex, baseline stroke severity and site of intracranial occlusion.

**Results:** Of the 1538 patients included in this analysis, 1146(74.5%)[591: tenecteplase, 555 alteplase] presented within 3 hours vs. 392 (25.5%)[196: TNK, 196 alteplase] who presented within 3-4.5 hours of symptom onset. Baseline patient characteristics in the 0-3 hour versus 3-4.5-hour time window were similar, except patients in the 3-to-4.5-hour window had lower median baseline NIHSS (10 vs 7 respectively) and lower proportion of patients with large vessel occlusion on baseline CT Angiography (26.9% vs 18.7% respectively). Type of thrombolytic agent (tenecteplase vs. alteplase) did not modify the association between ONT(p_interaction_ = 0.161) or DNT(p_interaction_ = 0.972) and primary clinical outcome. Irrespective of the thrombolytic agent used, each 30-min reduction in ONT was associated with a 1.8% increase while every 10 min reduction in DNT was associated with a 0.2% increase in the probability of achieving 90-day mRS 0-1 respectively.

**Conclusion:** The effect of time to tenecteplase administration on clinical outcomes is like that of alteplase, with faster administration resulting in better clinical outcomes.

**Key points:** *Question:* In patients with acute ischemic stroke, does the effect of time to thrombolysis on clinical outcomes differ with tenecteplase vs. alteplase administration?

*Findings:* In this analysis from the alteplase compared to tenecteplase (AcT) trial, a pragmatic, registry linked, phase 3 randomized controlled trial, each 30-min reduction in stroke onset to thrombolysis start time was associated with a 1.8% increase in the probability of achieving excellent functional outcome, which means that for every 30-minute reduction in onset to needle time two more of a 100 people achieved an excellent outcome. This effect was not modified by type of thrombolytic used (alteplase versus tenecteplase)

*Meaning:* The effect of time to tenecteplase administration on clinical outcomes is like that of alteplase, with faster administration resulting in better clinical outcomes.

## Introduction

Current American Heart Association/Stroke guidelines recommend the use of alteplase for intravenous thrombolysis (IVT) in patients with acute ischemic stroke presenting within 4.5 hours of symptom onset^1–3^. Tenecteplase, a second generation modified recombinant tissue type plasminogen activator, is more fibrin specific, has a longer half-life and depletes less systemic fibrinogen than alteplase^4^. Tenecteplase is also easier to administer as a single bolus over 5-10 seconds when compared to alteplase (bolus + 1 hour infusion), potentially facilitating faster treatment and transport of acute stroke patients within and between hospitals. Multiple phase 2 studies^4, 5^ and the recent large phase III Alteplase Compared to Tenecteplase (AcT) and Tenecteplase versus alteplase in acute ischemic cerebrovascular events (TRACE-2) randomized controlled trials show that intravenous tenecteplase (0.25mg/kg) is non-inferior to alteplase (0.9mg/kg) for IVT within 4.5 hours of symptom onset^6, 7^. These results are therefore supporting a transition to tenecteplase as the thrombolytic agent of choice for treating acute ischemic stroke.

Although the effect of time to treatment with alteplase on clinical outcomes is well known, with the phrase “Time is Brain” having become a key messaging strategy for acute stroke thrombolysis, the nature of this relationship in patients administered tenecteplase has yet to be described^8^. Moreover, with regulatory approval for alteplase restricted to patients presenting within 3 hours of acute ischemic stroke onset, the effect of thrombolysis with tenecteplase vs. alteplase in acute ischemic stroke patients presenting beyond 3 hours is also not understood well^9, 10^. The aim of this study was therefore to present the effect of thrombolysis with tenecteplase vs. alteplase on clinical outcomes in the early (0-3-hour time window) vs. the late (3-4.5-hour time window) presenting patients enrolled in the AcT RCT. In addition, the study also seeks to understand if the effect of time to treatment on clinical outcomes in patients receiving tenecteplase is any different from that of alteplase.

## Methods

### Study design and patient selection

The ACT trial (clinicaltrials.gov NCT03889249) was an investigator-initiated, large, pragmatic, multicenter, open-label, registry-linked, randomized, controlled, non-inferiority trial, with blinded end-point assessment (PROBE), comparing tenecteplase to alteplase in patients presenting with acute ischemic stroke. Inclusion and exclusion criteria were informed by the Canadian Stroke Best Practice Recommendations (CSBPR 2018)^11^, with patients eligible for inclusion if they were aged 18 years or older, with a diagnosis of acute ischemic stroke causing disabling neurological deficits, presenting within 4·5 hours of symptom onset, and eligible for thrombolysis. The trial enrolled 1600 patients from a total of 22 primary and comprehensive stroke centers across Canada between December 2019 and January 2022^6^. The 22 participating sites also participated in either the QuiCR (Quality Improvement and Clinical Research) or OPTIMISE (Optimizing Patient Treatment in Major Ischemic Stroke with EVT) acute stroke registries^12^. These registries provided workflow data to the trial. Workflow data included time of stroke symptom onset, hospital arrival and IVT administration. Stroke symptom onset was defined as time of witnessed onset of stroke symptoms. Door/Hospital arrival was defined as arrival at the emergency department of the thrombolysis-capable hospital. Needle time was defined as time of start of IVT. Only patients who received any dose of IVT and with symptom onset within 4.5 hours were included in the current analysis. The trial used deferred consent procedures wherever approved by local research ethics boards. Patients or their legal representatives were asked to provide informed consent as soon as possible after treatment, within 7 days of randomization or before discharge, whichever was earlier^6, 12, 13^.

### Outcomes

The primary outcome was the proportion of patients who achieved a score of 0 or 1 on the modified Rankin scale (mRS, an ordinal 7-point scale to assess disability after stroke with a score of 0 being not disability at all and 6 being death) at 90 days, up to 120 days after randomization. Secondary outcomes were mRS 0-2 at 90 days, ordinal mRS scale at 90 days, length of hospital stay and return to baseline function. These outcomes were all assessed by blinded assessors. Key safety outcomes were symptomatic intracerebral hemorrhage, extracranial bleeding requiring blood transfusion within 24 hours and 90-day all-cause mortality ^14^.

## Statistical Analyses

Data on baseline characteristics were summarized using frequencies and proportions for categorical variables and means (or medians) with standard deviations (or interquartile range) as appropriate for continuous variables. Differences in baseline characteristics amongst patients presenting early i.e., with onset to needle time (0-3 hours vs. late (3-4.5 hours) were analyzed using chi square test for categorical outcomes and t-test or Wilcoxon rank-sum test for continuous outcomes. Differences in primary, secondary and safety outcomes in patients receiving tenecteplase vs. alteplase presenting early and those presenting late are reported separately using unadjusted and adjusted analysis (adjusted for age, sex, baseline stroke severity measured using the National Institute of Health Stroke Scale [NIHSS]) and site of intracranial occlusion, with “enrolling site” as the random-effects variable to account for clustering of data within sites.

To model the association between time to treatment and each outcome, two clinically relevant interval times were used, namely, symptom onset to start of IVT (onset to needle time, ONT) and hospital arrival to start of IVT time (door to needle time, DNT) in two separate models. Patients with in-hospital stroke were excluded from the analysis of door to needle time as these patients arrived in the hospital before stroke onset and for reasons other than a suspected acute stroke. The functional form of the association between onset to needle time (ONT) and primary clinical outcome was examined using restricted cubic splines to evaluate potential nonlinear relationships between interval times and outcomes in patients receiving tenecteplase vs. alteplase, with 3 knots at 10^th^, 50^th^ and 90^th^ percentiles ^15^. The adjusted effect of onset to needle time on the primary outcome was also examined using mixed-effects logistic regression that included a time*treatment type multiplicative interaction term with age, sex, baseline NIHSS, and site of intracranial occlusion as fixed-effects variables, and “enrolling site” as the random-effects variable. Similar analyses as above was repeated with door to needle time (DNT) as independent variable instead of onset to needle time. Predicted probabilities of primary clinical outcome as a function of each of these interval times (onset to needle time and door to needle time) were estimated using mixed effects logistic regression models for each time interval. Similar analyses were attempted for all other outcomes. All analyses were considered exploratory. All reported p-values were two-sided, with p<0.05 considered statistically significant. Statistical analysis was performed using Stata/MP version 16.0 (StataCorp LP)^16^

## Results

Of the 1600 patients randomized in the AcT randomized controlled trial, 1577 (98.6%) consented to study participation. Of these, 14 patients who did not receive IV thrombolysis and 25 patients who received IV thrombolysis beyond 4.5 hours of symptom onset were excluded from the analysis (**Supplemental Figure 1**; Study Flowchart). Of the remaining 1538 included patients, 1146(74.5%, 591: tenecteplase arm and 555: alteplase arm) were in the 0–3-hour ONT window and 392 (25.5%, 196: tenecteplase arm and 196: alteplase arm) were in the 3-4.5-hour ONT window. Baseline patient characteristics by the pre-specified time windows and stratified by treatment type (tenecteplase vs. alteplase) are shown in **Table 1 and Supplemental Table 1** respectively. Patients in the 3-4.5-hour window had lower median baseline NIHSS (10 versus 7 respectively) and lower proportion of patients with large vessel occlusion (26.9% vs 18.7% respectively). In patients who underwent EVT, median CT to arterial puncture times and arterial puncture to reperfusion times were 9 mins and 6 mins faster in the 3-4.5-hour window vs. 0–3-hour time window (**Table 1**).

**Table 1.**
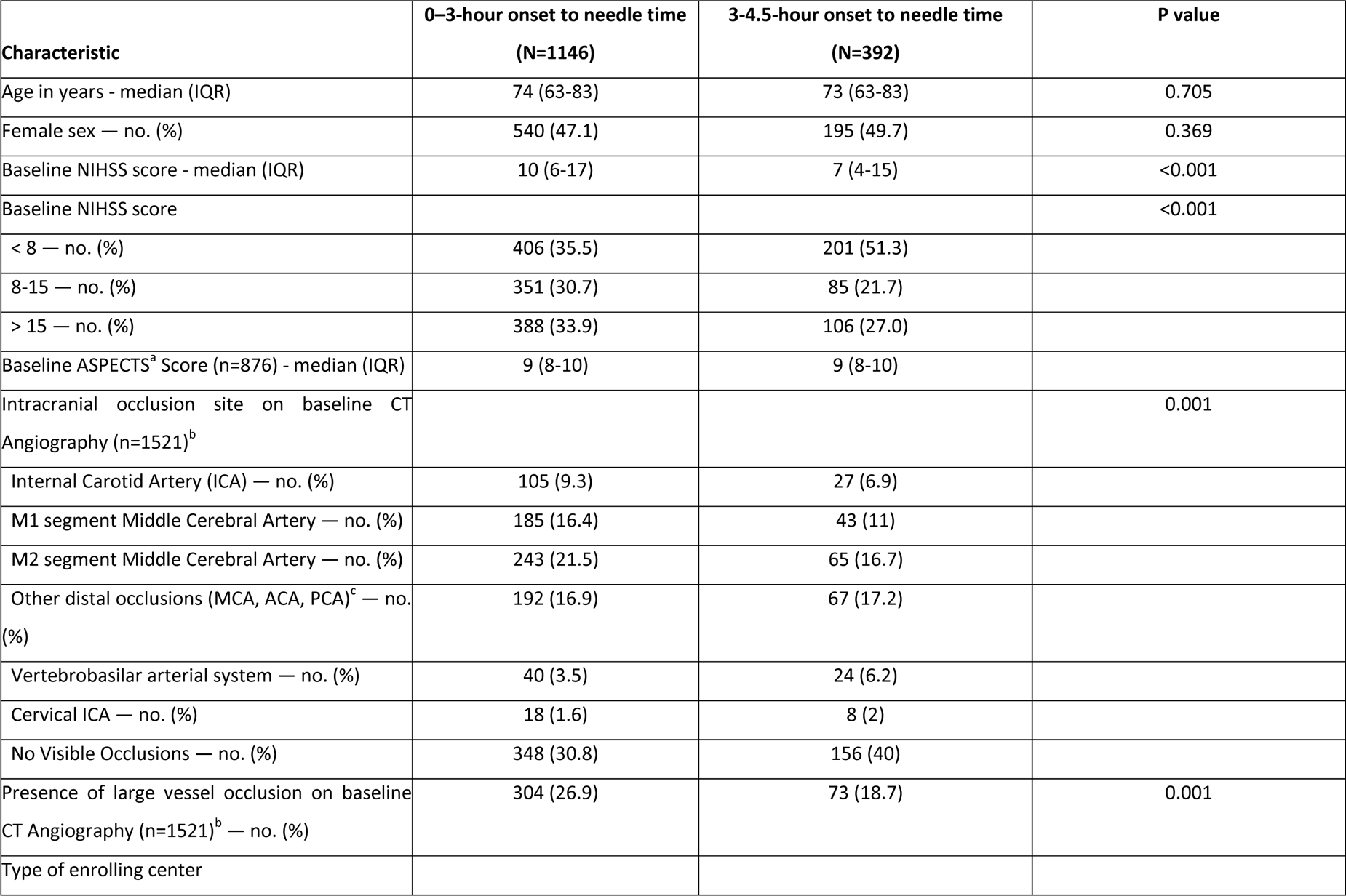
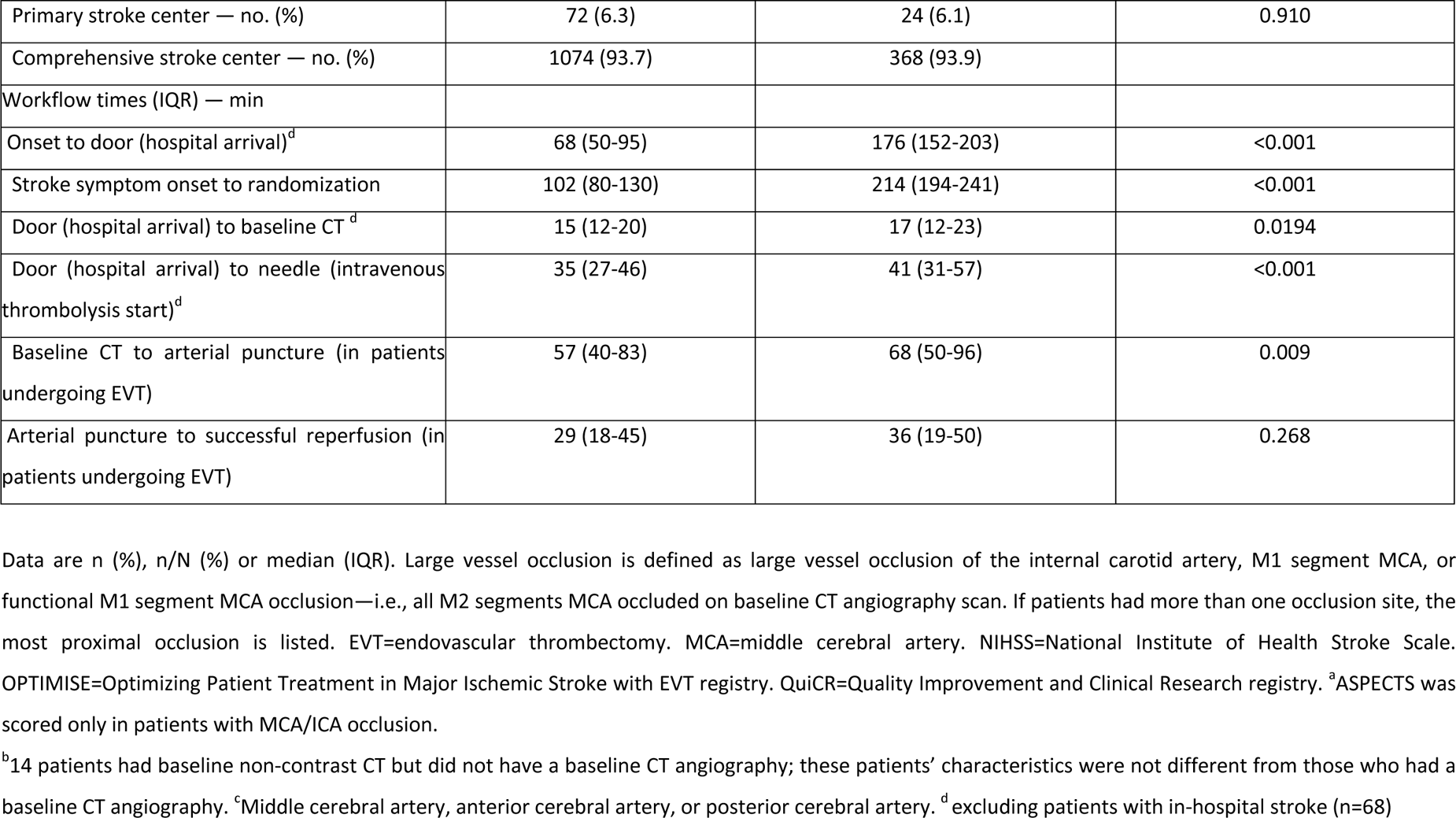
Baseline characteristics in patients treated within 0-3 hour and 3-4.5-hour windows.

| Characteristic | 0-3-hour onset to needle time<br>(N=1146) | 3-4.5-hour onset to needle time<br>(N=392) | P value |
| --- | --- | --- | --- |
| Age in years - median (IQR) | 74 (63-83) | 73 (63-83) | 0.705 |
| Female sex — no. (%) | 540 (47.1) | 195 (49.7) | 0.369 |
| Baseline NIHSS score - median (IQR) | 10 (6-17) | 7 (4-15) | <0.001 |
| Baseline NIHSS score |  |  | <0.001 |
| < 8 — no. (%) | 406 (35.5) | 201 (51.3) |  |
| 8-15 — no. (%) | 351 (30.7) | 85 (21.7) |  |
| > 15 — no. (%) | 388 (33.9) | 106 (27.0) |  |
| Baseline ASPECTS<sup>a</sup> Score (n=876) - median (IQR) | 9 (8-10) | 9 (8-10) |  |
| Intracranial occlusion site on baseline CT Angiography (n=1521)<sup>b</sup> |  |  | 0.001 |
| Internal Carotid Artery (ICA) — no. (%) | 105 (9.3) | 27 (6.9) |  |
| M1 segment Middle Cerebral Artery — no. (%) | 185 (16.4) | 43 (11) |  |
| M2 segment Middle Cerebral Artery — no. (%) | 243 (21.5) | 65 (16.7) |  |
| Other distal occlusions (MCA, ACA, PCA)<sup>c</sup> — no. (%) | 192 (16.9) | 67 (17.2) |  |
| Vertebrobasilar arterial system — no. (%) | 40 (3.5) | 24 (6.2) |  |
| Cervical ICA — no. (%) | 18 (1.6) | 8 (2) |  |
| No Visible Occlusions — no. (%) | 348 (30.8) | 156 (40) |  |
| Presence of large vessel occlusion on baseline CT Angiography (n=1521)<sup>b</sup> — no. (%) | 304 (26.9) | 73 (18.7) | 0.001 |
| Type of enrolling center |  |  |  |
| Primary stroke center — no. (%) | 72 (6.3) | 24 (6.1) | 0.910 |
| Comprehensive stroke center — no. (%) | 1074 (93.7) | 368 (93.9) |  |
| Workflow times (IQR) — min |  |  |  |
| Onset to door (hospital arrival)<sup>d</sup> | 68 (50-95) | 176 (152-203) | <0.001 |
| Stroke symptom onset to randomization | 102 (80-130) | 214 (194-241) | <0.001 |
| Door (hospital arrival) to baseline CT<sup>d</sup> | 15 (12-20) | 17 (12-23) | 0.0194 |
| Door (hospital arrival) to needle (intravenous thrombolysis start)<sup>d</sup> | 35 (27-46) | 41 (31-57) | <0.001 |
| Baseline CT to arterial puncture (in patients undergoing EVT) | 57 (40-83) | 68 (50-96) | 0.009 |
| Arterial puncture to successful reperfusion (in patients undergoing EVT) | 29 (18-45) | 36 (19-50) | 0.268 |

Data are n (%), n/N (%) or median (IQR). Large vessel occlusion is defined as large vessel occlusion of the internal carotid artery, M1 segment MCA, or functional M1 segment MCA occlusion—i.e., all M2 segments MCA occluded on baseline CT angiography scan. If patients had more than one occlusion site, the most proximal occlusion is listed. EVT=endovascular thrombectomy. MCA=middle cerebral artery. NIHSS=National Institute of Health Stroke Scale. OPTIMISE=Optimizing Patient Treatment in Major Ischemic Stroke with EVT registry. QuiCR=Quality Improvement and Clinical Research registry. <sup>a</sup>ASPECTS was scored only in patients with MCA/ICA occlusion.
<sup>b</sup>14 patients had baseline non-contrast CT but did not have a baseline CT angiography; these patients' characteristics were not different from those who had a baseline CT angiography. <sup>c</sup>Middle cerebral artery, anterior cerebral artery, or posterior cerebral artery. <sup>d</sup>excluding patients with in-hospital stroke (n=68)

### Comparison by thrombolytic type (Tenecteplase vs. alteplase) in the 0–3-hour time window

In the 0–3-hour window, baseline patient characteristics and workflow times were similar in the two treatment arms (**Supplemental Table 1**). The primary outcome (90-day mRS 0–1) occurred in 216 (36.7%) of 591 patients in the tenecteplase arm and 198 (35.9%) of 555 patients in the alteplase arm (unadjusted odds ratio 1.03 [95% CI 0.81 to 1.31; **Table 2**). Secondary outcomes such as mRS 0-2 at 90 days, median mRS at 90days, return to baseline function and length of hospital stay were similar in both treatment arms (**Table 2**). Rate of 24h symptomatic intracerebral hemorrhage in the tenecteplase vs. alteplase arm (3.2% vs 3.1%) were not different (unadjusted odds ratio 1.23, 95% CI [1.05(0.54-2.04]). There were no meaningful differences in mortality or other safety outcomes (**Table 3, Figure 3A & B**). No meaningful differences were noted for any outcomes in adjusted analysis. (**Tables 2 and 3**).

**Table 2:** Efficacy outcomes in patients treated with intravenous tenecteplase vs. alteplase within the 0-3 hour and 3-4.5-hour time windows.

|  | 0–3-hour window |  |  |  | 3-4.5-hour window |  |  |  |
| --- | --- | --- | --- | --- | --- | --- | --- | --- |
| Primary end point | Tenecteplase group<br>(N=591) n,(%) | Alteplase group<br>(N=555) n,(%) | Odds Ratio<br>(unadjusted) | Odds Ratio<br>(Adjusted)<sup>a</sup> | Tenecteplase group<br>(N=196) | Alteplase group<br>(N=196) | Odds Ratio<br>(unadjusted) | Odds Ratio<br>(Adjusted)<sup>a</sup> |
| modified Rankin Score 0-1 at 90 days — no. (%) | 216 (36.7) | 198 (35.9) | 1.03 (0.81-1.31) | 1.03 (0.80-1.34) | 76 (38.9) | 61 (31.3) | 1.40 (0.92-2.13) | 1.47 (0.94-1.2.29) |
| Secondary end points |  |  |  |  |  |  |  |  |
| modified Rankin Score 0-2 at 90 days — no. (%) | 331 (56.2) | 313 (56.8) | 0.97 (0.77-1.23) | 0.94 (0.72-1.22) | 111 (56.9) | 102 (52.3) | 1.20 (0.80-1.79) | 1.34 (0.84-2.12) |
| modified Rankin Score at 90 days - median (IQR) | 2 (1-4) | 2 (1-4) | 0.97 (0.79-1.19)<sup>b</sup> | 0.98 (0.79-1.20)<sup>b</sup> | 2 (1-4) | 2 (1-4) | 0.79 (0.55-1.21) | 0.76 (0.53-1.08)<sup>b</sup> |
| Return to baseline function — no. (%) | 160 (29.6) | 155 (29.9) | 0.99 (0.75-1.28) | 1.01 (0.76-1.33) | 54 (29.8) | 37 (20.9) | 1.60 (0.99-2.60) | 1.68 (0.98-2.88) |
| Endovascular Thrombectomy Utilization — no. (%) | 215 (36.4) | 197 (35.5) | 1.03 (0.81-1.32) | 0.95 (0.65-1.39) | 36 (18.4) | 45 (22.9) | 0.75 (0.46-1.23) | 0.97 (0.49-1.93) |
| Length of hospital stay - median (IQR) | 5 (2-11) | 5 (3-11) | 0.98 (0.80-1.21)<sup>b</sup> | 1.03 (0.83-1.27)<sup>b</sup> | 5 (3-10) | 5 (3-12) | 0.94 (0.66-1.34)<sup>b</sup> | 0.40 (0.07-2.32)<sup>b</sup> |

Data are n (%), median (IQR), mean (SD), or effect estimate with 95% CI in parentheses. <sup>a</sup>Adjusted for age, sex, baseline stroke severity and occlusion site as fixed- effects variables, and site as a random effects variable. <sup>b</sup>Common odds ratio is the odds ratio for a unit increase in the modified Rankin scale score for tenecteplase vs alteplase.

**Table 3:** Safety Outcomes in patients treated with intravenous tenecteplase vs. alteplase within 0-3 hour and 3-4.5-hour time windows.

|  | 0–3-hour window |  |  |  | 3-4.5-hour window |  |  |  |
| --- | --- | --- | --- | --- | --- | --- | --- | --- |
| End points | Tenecteplase group (N=591) | Alteplase group (N=555) | Odds Ratio (unadjusted) | Odds Ratio (adjusted) | Tenecteplase group (N=196) | Alteplase group (N=196) | Odds Ratio (unadjusted) | Odds Ratio (unadjusted) |
| Death within 90 days — no. (%) | 83 (15.1) | 88 (14.9) | 0.99 (0.71-1.37) | 1.06 (0.74-1.50) | 31 (15.9) | 33 (16.9) | 0.93 (0.54-1.58) | 0.86 (0.47-1.58) |
| Symptomatic intracranial hemorrhage — no. (%) | 19 (3.2) | 17 (3.1) | 1.05 (0.54-2.04) | 1.14 (0.57-2.27) | 7 (3.6) | 7 (3.6) | 1.0 (0.34-2.90) | 1.0 (0.33-2.98) |
| Peripheral Bleeding requiring blood transfusions — no. (%) | 6 (1) | 4 (0.7) | 1.41 (0.39-5.03) | 1.53 (0.41-5.80) | 0 (0) | 2 (1.0) | -- | -- |
| Orolingual angioedema — no. (%) | 6 (1) | 8 (1.4) | 0.70 (0.24-2.03) | 0.69 (0.23-2.08) | 3 (1.5) | 0 (0) | -- | -- |
| Others — no. (%) | 66 (11.2) | 45 (8.1) | 1.42 (0.95-2.12) | 1.44 (0.95-2.17) | 13 (6.6) | 23 (11.7) | 0.53 (0.26-1.08) | 0.56 (0.26-1.19) |
| Imaging identified hemorrhage — no. (%) | 117 (19.9) | 113 (20.6) | 0.95 (0.71-1.27) | 0.97 (0.71-1.31) | 34 (17.5) | 42 (21.5) | 0.77 (0.46-1.28) | 0.78 (0.46-1.35) |
| Subarachnoid hemorrhage (SAH) — no. (%) | 46 (7.8) | 41 (7.5) | 1.05 (0.70-1.58) | 1.07 (0.68-1.70) | 5 (2.6) | 11 (5.6) | 0.44 (0.15-1.29) | 0.41 (0.14-1.22) |
| Subdural hemorrhage (SDH) — no. (%) | 2 (0.3) | 4 (0.7) | 0.46 (0.08-2.53) | 0.39 (0.07-2.24) | 0 (0) | 1 (0.5) | -- | -- |
| Intraventricular | 18 (3.1) | 11 (2.0) | 1.53 (0.72-3.28) | 1.57 (0.72-3.38) | 6 (3.1) | 6 (3.1) | 1.00 (0.32-3.17) | 0.94 (0.29-2.97) |

| hemorrhage (IVH) —<br>no. (%) |  |  |  |  |  |  |  |  |
| --- | --- | --- | --- | --- | --- | --- | --- | --- |
| HI1 (scattered small<br>petechiae) | 9 (1.5) | 17 (3.1) | 0.48 (0.21-1.09) | 0.48 (0.20-1.01) | 9 (4.6) | 7 (3.6) | 1.30 (0.47-3.58) | 1.27 (0.43-3.72) |
| HI2 (confluent<br>petechiae) | 51 (8.7) | 44 (8.0) | 1.08 (0.71-1.65) | 1.09 (0.71-1.68) | 9 (4.6) | 22 (11.3) | 0.38 (0.17-0.85) | 0.37 (0.16-0.84) |
| PH1 (hematoma<br>occupying <30% of<br>infarct with no<br>substantive mass<br>effect) | 21 (3.6) | 15 (2.7) | 1.31 (0.67-2.57) | 1.38 (0.68-2.78) | 7 (3.6) | 4 (2.0) | 1.70 (0.51-6.20) | 1.71 (0.49-5.97) |
| PH2 (hematoma<br>occupying ≥30% of<br>infarct with obvious<br>mass effect) | 17 (2.9) | 13(2.4) | 1.22 (0.59-2.54) | 1.25 (0.59-2.62) | 5 (2.6) | 4 (2.1) | 0.8 (0.21-3.02) | 0.72 (0.18-2.83) |
| Remote PH1<sup>a</sup> | 5 (0.8) | 6 (1.1) | 0.77 (0.23-2.53) | 0.76 (0.23-2.58) | 1 (0.5) | 3 (1.5) | 0.33 (0.03-3.21) | 0.27 (0.03-2.68) |
| Remote PH2<sup>b</sup> | 1 (0.2) | 2 (0.4) | 0.56 (0.04-5.13) | 0.46 (0.04-5.15) | 1 (0.5) | 1 (0.5) | 1.00 (0.06-16.2) | 0.79 (0.03-16.2) |

Data are n (%) or risk ratio with 95% CI in parentheses. Imaging-identified intracranial hemorrhages were assessed in a central core laboratory in a blinded manner and classified using the Heidelberg classification. <sup>a</sup>Remote parenchymal hematoma type 1 was defined as hematoma outside the infarcted tissue with no substantive mass effect. <sup>b</sup>Remote parenchymal hematoma type 2 was defined as hematoma outside the infarcted tissue, with obvious mass effect.

### Comparison by thrombolytic type in the 3–4.5-hour time window

In the 3-4.5-hour window, baseline characteristics were similar in the two treatment arms, except that higher proportion of patients in the tenecteplase arm had baseline NIHSS <=8 (57.1% vs 45.4%), and fewer patients had large vessel occlusion on baseline CT Angiography (14.8% vs 22.6%) as compared to alteplase. (**Supplemental Table 1**). The primary outcome (90-day mRS 0–1) occurred in 76 (38.9%) of 196 patients in the tenecteplase arm and 61 (31.3%) of 196 patients in the alteplase arm (unadjusted odds ratio 1.40 [95% CI 0.92 to 2.13]; **Table 2**). Secondary outcomes such as mRS 0-2 at 90 days, median mRS at 90days, return to baseline function and length of hospital stay were similar in both treatment arms (**Table 2**). Rate of 24h symptomatic intracerebral hemorrhage in the tenecteplase vs. alteplase arm (3.6% vs 3.6%) were not different [unadjusted odds ratio 1.0 (0.34-2.90)]. There were no meaningful differences in mortality or other safety outcomes (**Table 3, Figure 3A & B**). No meaningful differences were noted for any outcomes in adjusted analysis. (**Tables 2 and 3**).

### Association between Time to Treatment (OTN and DTN) vs. outcomes

The restricted cubic spline models of the association between ONT vs. primary outcome (mRS 0-1 at 90 days) stratified by thrombolytic type (tenecteplase vs. alteplase) shows a linear relationship was most appropriate for modeling the association between ONT and mRS0-1 (**Figure 1A**). Results from the adjusted analyses using mixed effects logistic regression show that the association between ONT and 90-day mRS 0-1 was not modified by type of thrombolytic administered (tenecteplase vs. alteplase) (p value for interaction 0.161) (**Figure 1B**). The probability of excellent functional outcome (mRS 0-1 at 90 days) increased by 1.8% for every 30-min reduction in onset to needle time (**Supplemental Figure 2**). There was no evidence of treatment effect modification of the relationship between ONT vs. any other outcome (**Supplemental Table 2, Figure 2A and 2B**).

**Figure 1:**
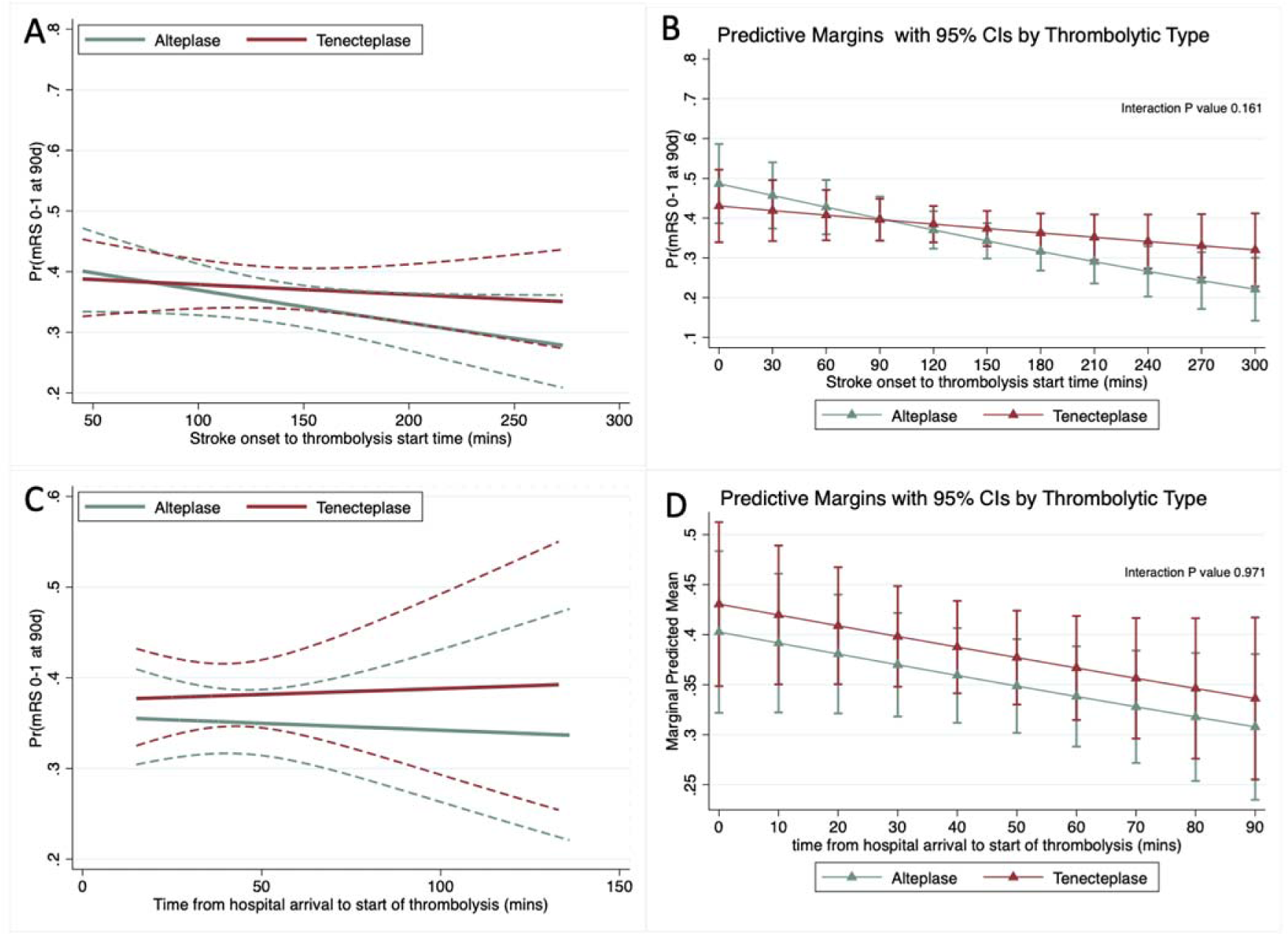
Relationship between onset to needle time and mRS 0-1 at 90-120 days stratified by thrombolytic type using restricted cubic spline model (A) and mixed effects logistic regression model (B) adjusted for age, sex, stroke severity as measured using the National Institute of Health Stroke Scale (NIHSS) and intracranial occlusion site as fixed effects variables and “enrolling site” as the random effects variable (error bars indicate 95% CI). Relation between door to needle time and mRS 0-1 at 90-120 days stratified by thrombolytic type using restricted cubic spline model (C) and mixed effects logistic regression model (D) adjusted for age, sex, stroke severity and occlusion site with site as random effects (error bars indicate 95% CI)

The restricted cubic spline model of the relationship between DNT and primary clinical outcome (mRS 0-1 at 90 days) stratified by thrombolytic type (tenecteplase vs. alteplase) was also noted to be linear (**Figure 1C**). Similar to analyses with ONT above, with a mixed effects logistic regression model, the relationship between DNT and 90-day mRS 0-1 was not modified by type of thrombolytic administered (tenecteplase vs. alteplase) (p value for interaction 0.971) (**Figure 1D**). The probability of excellent functional outcome (mRS 0-1 at 90 days) decreased by 0.2% for every 10-min delay in door to needle time (**Supplemental Figure 2**). There was no evidence of treatment effect modification of the relationship between DNT vs. any other outcome (**Supplemental Table 2, Figure 2C and 2D**).

**Figure 2:**
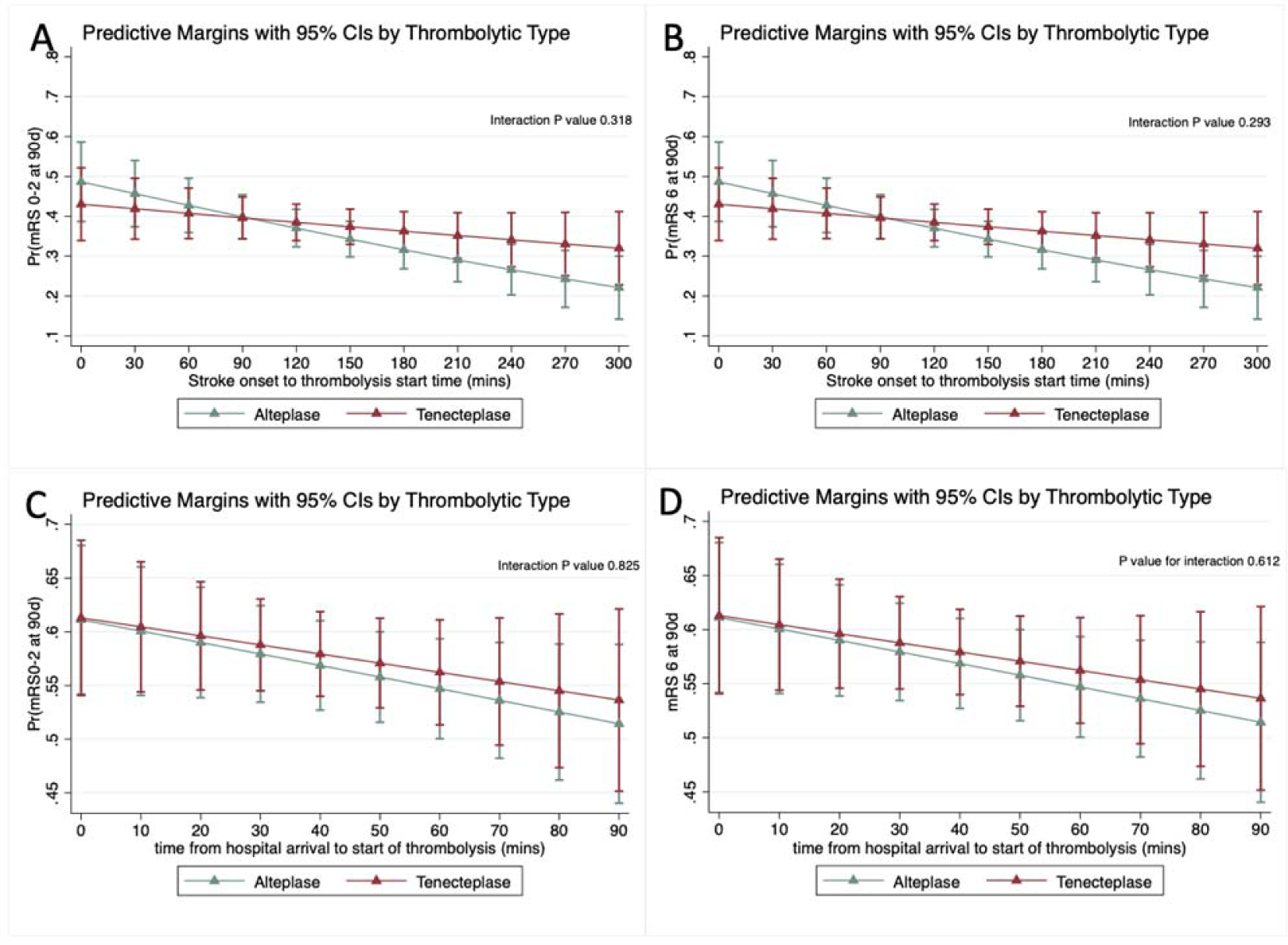
Relationship between onset to needle time and mRS 0-2 at 90-120d days (A) and death at 90-120d (B) using mixed effects logistic regression model adjusted for age, sex, stroke severity as measured using the National Institute of Health Stroke Scale (NIHSS) and intracranial occlusion site as fixed effects variables and “enrolling site” as the random effects variable (error bars indicate 95% CI). Relation between door to needle time and mRS 0-2 at 90-120d days (C) and death at 90-120d (D) stratified by thrombolytic type using mixed effects logistic regression model adjusted for age, sex, stroke severity and occlusion site with site as random effects (error bars indicate 95% CI)

**Figure 3:**
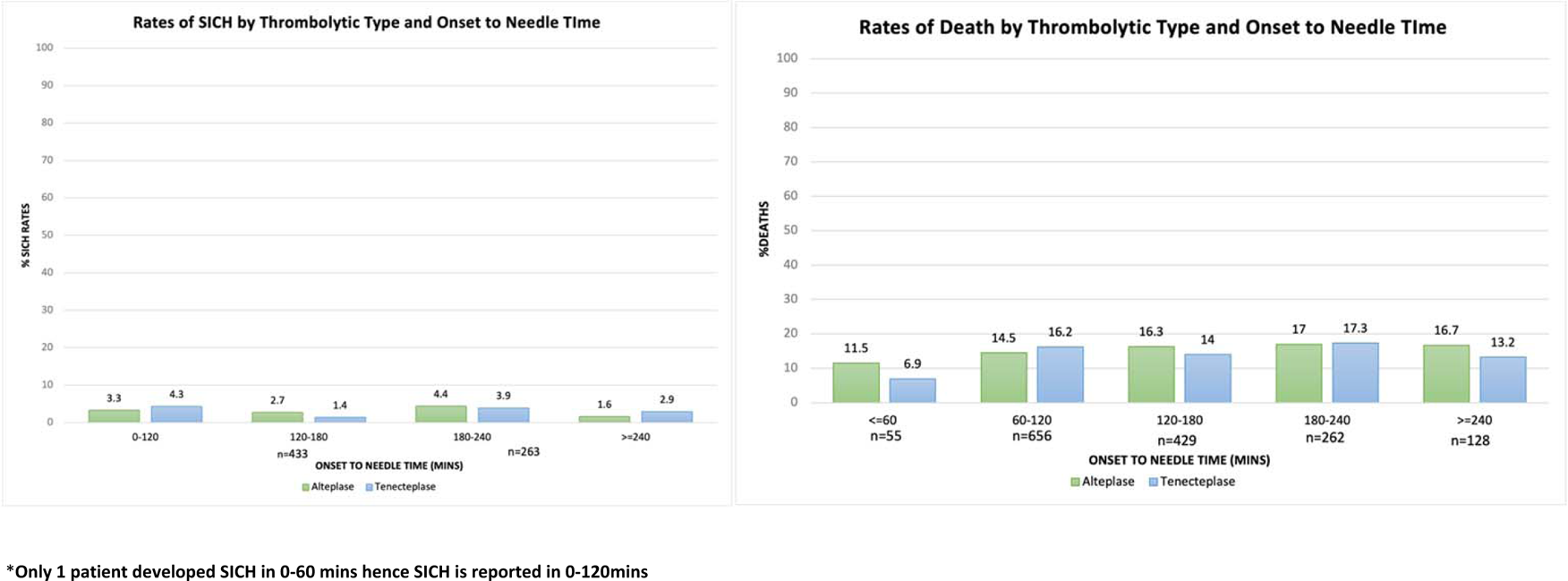
Proportion of safety outcomes [mortality (A), symptomatic intracerebral hemorrhage (B)] stratified by thrombolytic type across symptom onset to needle time categories.

## Discussion

This analysis from the recent phase 3 AcT randomized controlled trial provides supportive evidence that intravenous tenecteplase has similar effect on clinical outcomes as alteplase not just in patients presenting early (within 3 hours of stroke symptom onset) but also in those presenting late (3-4.5 hours)^6^. This analysis also shows that the detrimental effect of delay in thrombolysis on clinical outcomes in patients acute ischemic stroke is also similar with tenecteplase vs alteplase. Faster administration of tenecteplase, like alteplase, improves the chances of achieving better clinical outcomes in patients with acute ischemic stroke treated within 4.5 hours. These results also support the underlying principle of the AcT trial, that the decision to thrombolyse in an acute stroke setting can be made by pragmatic imaging criteria. Ultimately, acting rapidly and effectively on that information is what impacts patient outcomes.

Tenecteplase, a newer generation thrombolytic, is the standard of care in cardiology to treat patients with ST-elevation myocardial infarction who do not have immediate access to primary percutaneous intervention (PCI)^17^. In select patients with acute ischemic stroke, some phase 2 studies had shown increased reperfusion with tenecteplase vs. alteplase. ^5, 18, 19^ The AcT trial was the first phase 3 RCT to compare intravenous tenecteplase to alteplase in all patients presenting with acute ischemic stroke within 4.5 hours from symptom onset and eligible for thrombolysis with alteplase.^20^ The trial showed that tenecteplase (0.25mg/kg) was non inferior to alteplase (0.9mg/kg) in treatment of acute ischemic stroke (unadjusted risk difference 2.1% [95%CI - 2.6 to 6.9]) and comparable for all other secondary and safety outcomes ^6^. These results have changed Canadian, European and Australian guidelines, all of which now recommend use of tenecteplase for acute ischemic stroke within 4.5 hour of symptoms onset^20–22^. Unlike clinical practice guidelines, regulatory approval for intravenous alteplase use in patients with acute ischemic stroke is restricted to patients presenting within 3 hours of stroke symptom onset^9, 10^. This key secondary analyses from the AcT trial aims to therefore present in detail the effect of tenecteplase vs. alteplase in the early vs. late (3-4.5 hour) presenters while also assessing if the well-known relationship of time to thrombolysis on outcomes with alteplase is also seen with tenecteplase.

The study has several strengths and implications. One of the strengths is the use of a large sample size from the AcT RCT trial, which provides a robust representation of patients with acute ischemic stroke who were treated with either alteplase or tenecteplase. Additionally, the study used mixed effects logistic regression to adjust for potential confounding factors, which increases the accuracy of the results. These results have several important implications for clinical practice. First, the findings suggest that the association between time to thrombolysis treatment and clinical outcomes is similar for both alteplase and tenecteplase. This is important because tenecteplase has been shown to be non-inferior to alteplase in treating acute ischemic stroke within 4.5 hours of symptom onset. Second, the study reinforces the importance of reducing the time to thrombolysis treatment for patients with acute ischemic stroke. Specifically, the study found that each 30-minute reduction in ONT was associated with a 1.8% increase in the probability of achieving mRS 0-1 at 90 days, while every 10-minute reduction in DNT was associated with a 0.2% increase in the same outcome. These findings highlight the need for streamlined processes and protocols to ensure prompt and efficient thrombolysis treatment for patients with acute ischemic stroke. Overall, the study provides valuable insights into the association between time to thrombolysis treatment and clinical outcomes in patients with acute ischemic stroke treated with either alteplase or tenecteplase.

This study has some limitations. The AcT trial did not stratify randomization based on time from stroke onset to thrombolysis. Although adjusted analysis was used to mitigate the effect of any differences in baseline characteristics on clinical outcomes, such analysis may not have fully addressed any differences due to unmeasured confounders at baseline. Although time to treatment analysis was pre-specified in the AcT trial protocol, such analyses was considered exploratory. Finally, although the Canadian Stroke Best Practice recommendations are similar to current AHA/Stroke guidelines, we cannot discount the possibility of practice and health system differences in these two countries influencing some of these results.

In conclusion, this pre-specified but exploratory analysis from the large phase III AcT trial suggests that the effect of time to tenecteplase administration on clinical outcomes is like that of alteplase, with faster administration resulting in better clinical outcomes.

## Data Availability

Data will be made available upon reasonable request

## Acknowledgements

none

## Supplemental Material

**Supplemental Table 1:**
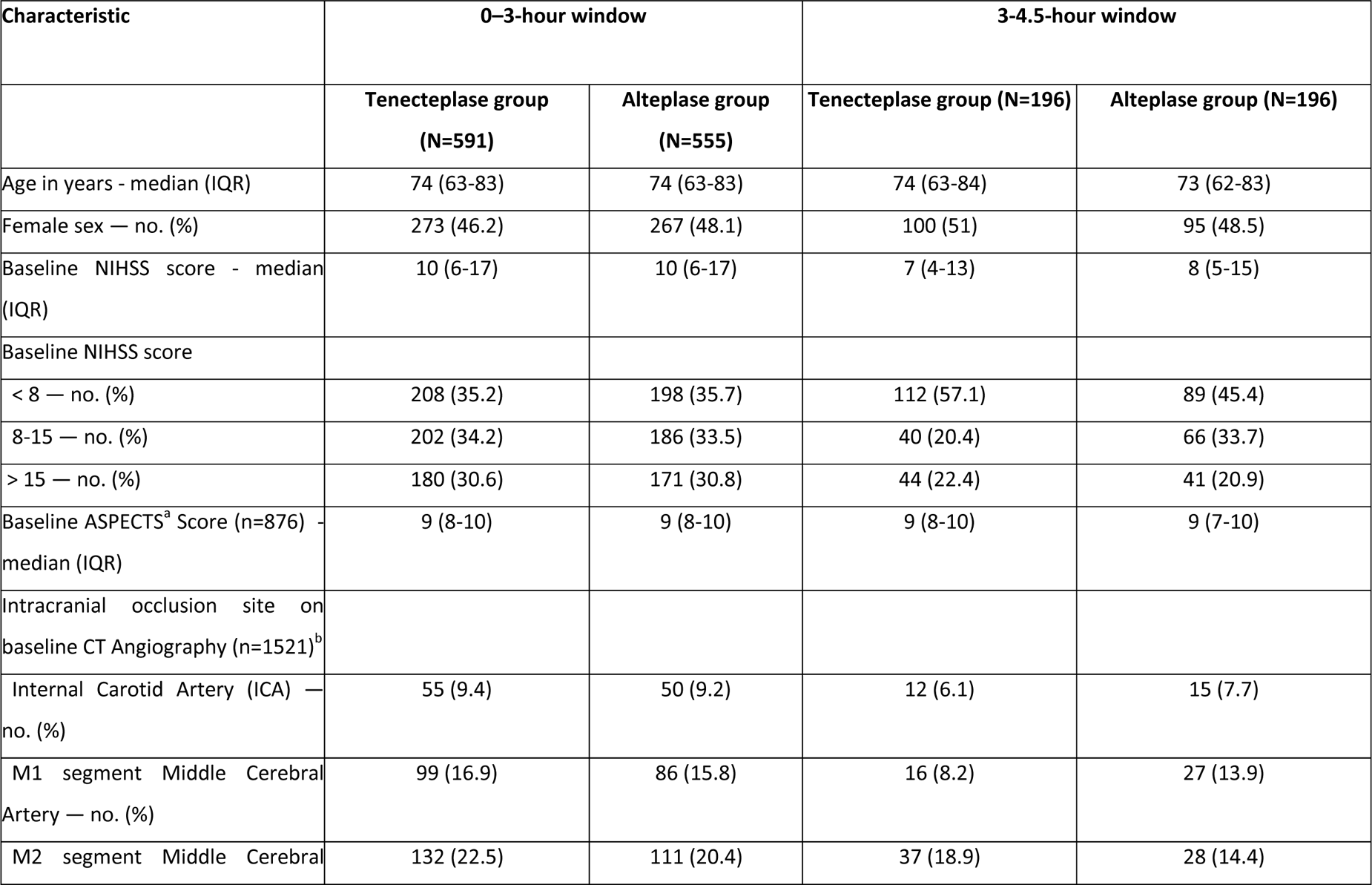
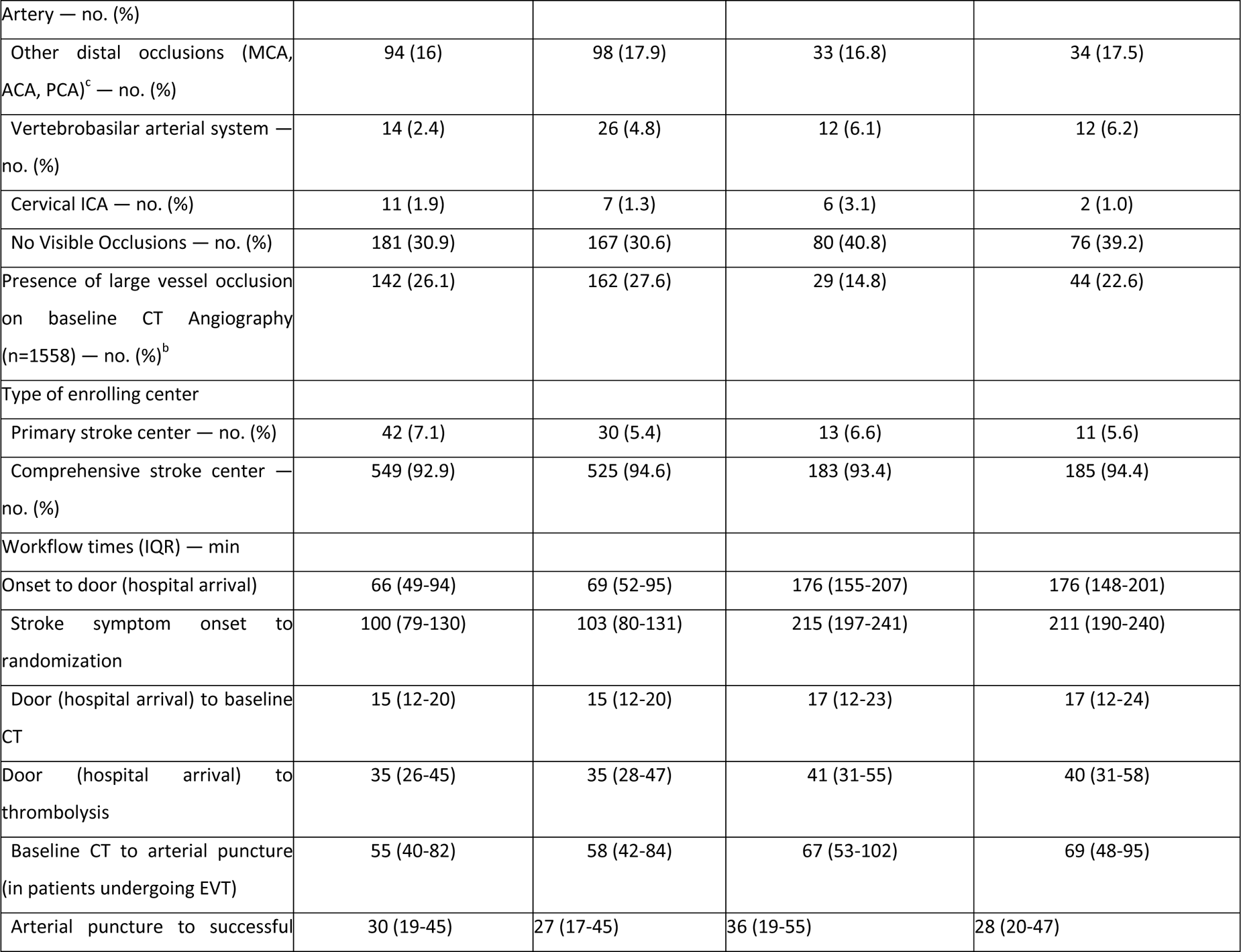
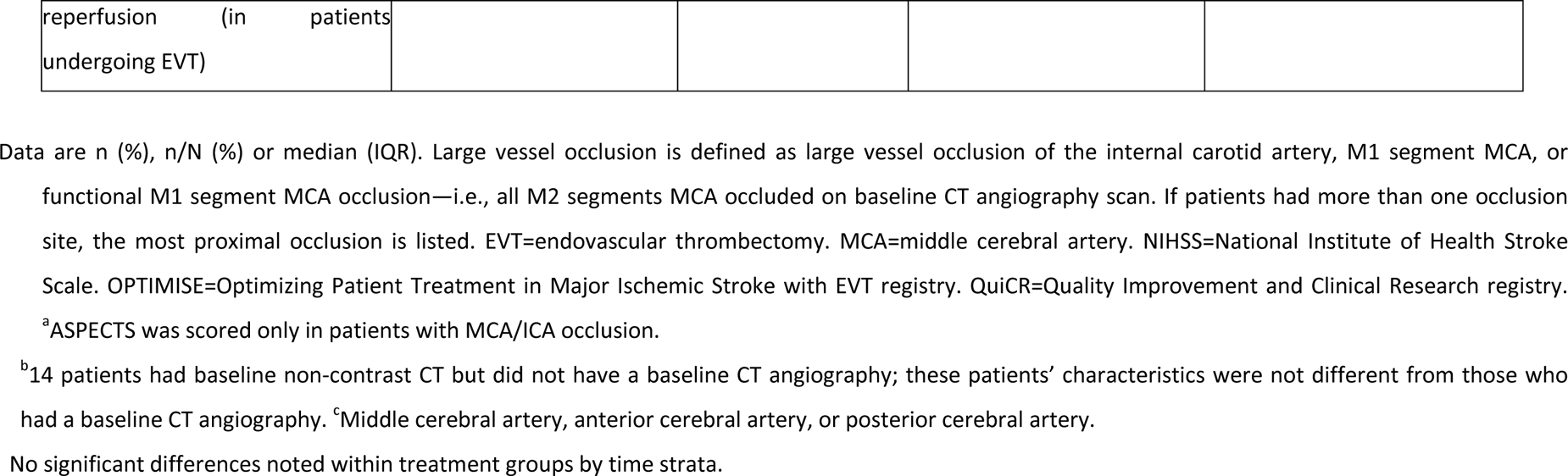
Baseline characteristics in patients treated within 0-3 hour and 3-4.5-hour window stratified by thrombolytic type.

**Supplemental Table 2.**
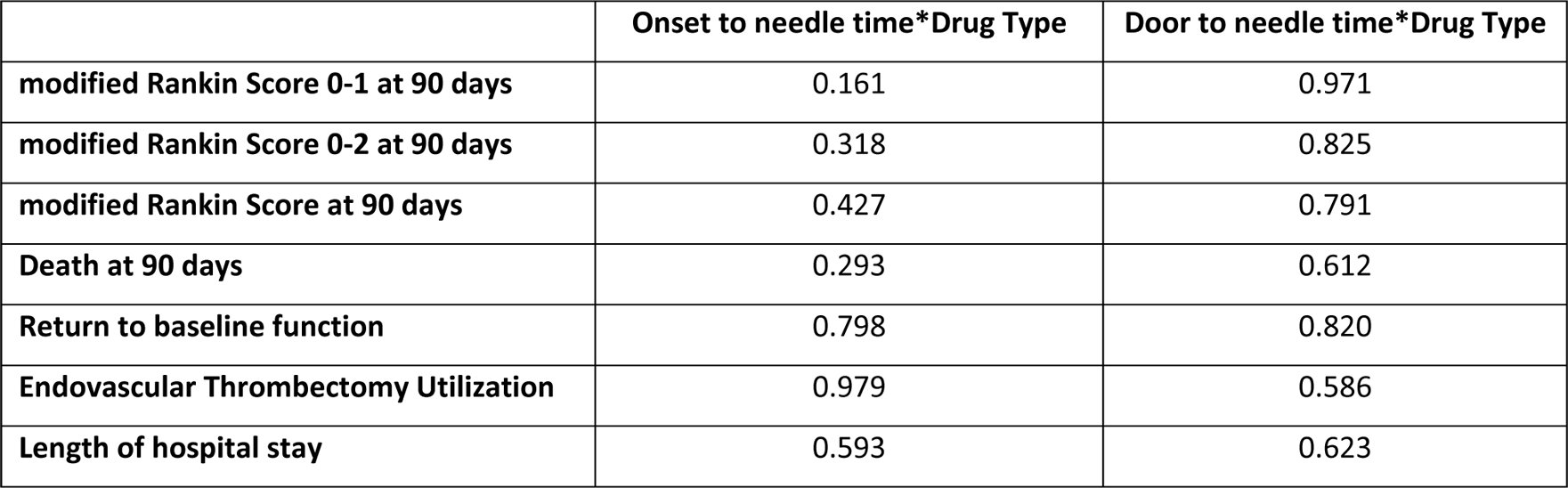
Interaction p values for multiplicative interaction term time*thrombolytic type using mixed effects logistic regression model adjusted for age, sex, baseline stroke severity as measured using the National Institute of Health Stroke Scale (NIHSS) and intracranial occlusion location as fixed effects variables with enrolling site as the random effects variable (error bars indicate 95% CI)

|  | Onset to needle time*Drug Type | Door to needle time*Drug Type |
| --- | --- | --- |
| modified Rankin Score 0-1 at 90 days | 0.161 | 0.971 |
| modified Rankin Score 0-2 at 90 days | 0.318 | 0.825 |
| modified Rankin Score at 90 days | 0.427 | 0.791 |
| Death at 90 days | 0.293 | 0.612 |
| Return to baseline function | 0.798 | 0.820 |
| Endovascular Thrombectomy Utilization | 0.979 | 0.586 |
| Length of hospital stay | 0.593 | 0.623 |

**Supplemental Figure 1:**
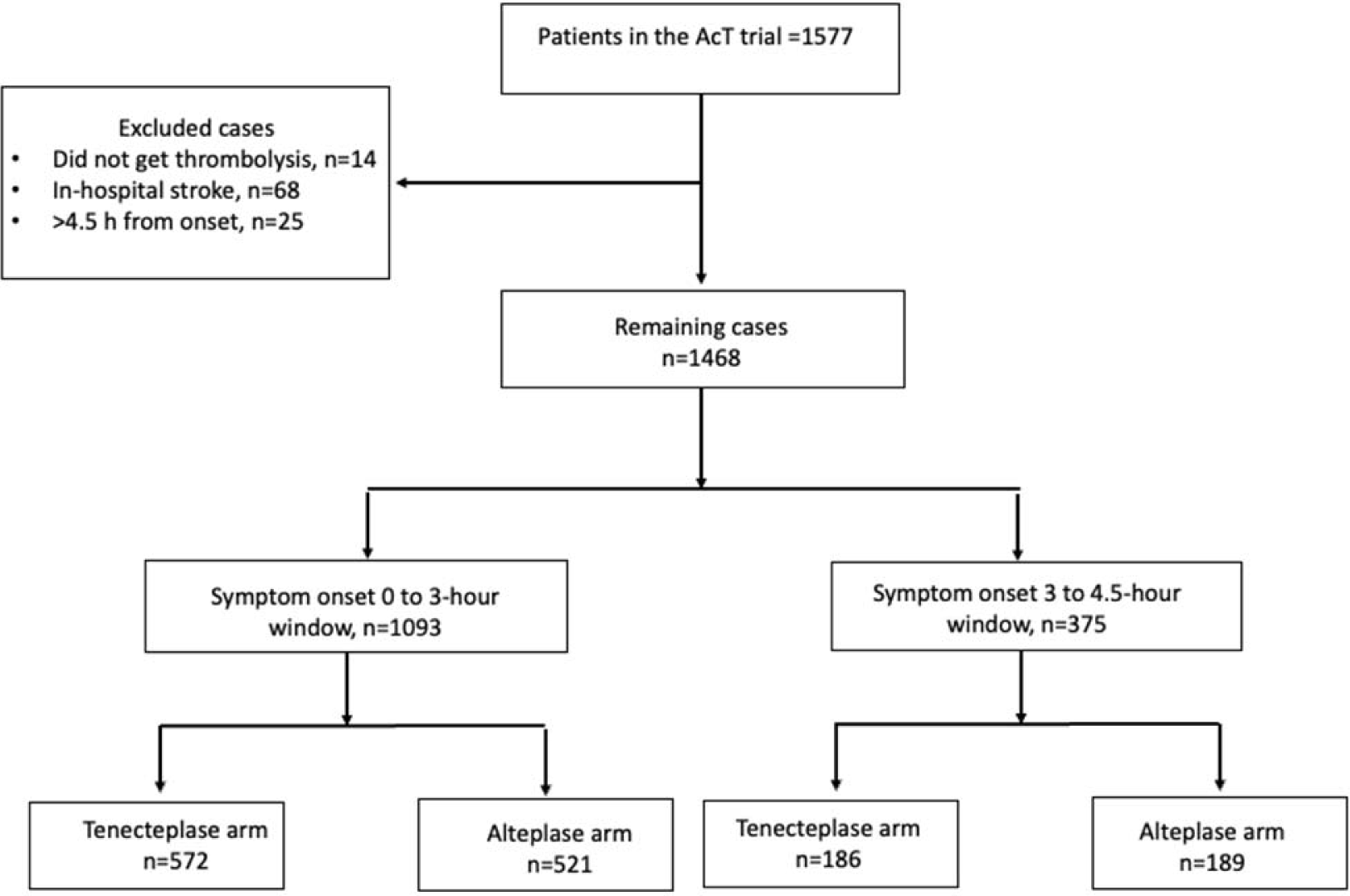
Study Flowchart

**Supplemental Figure 2:**
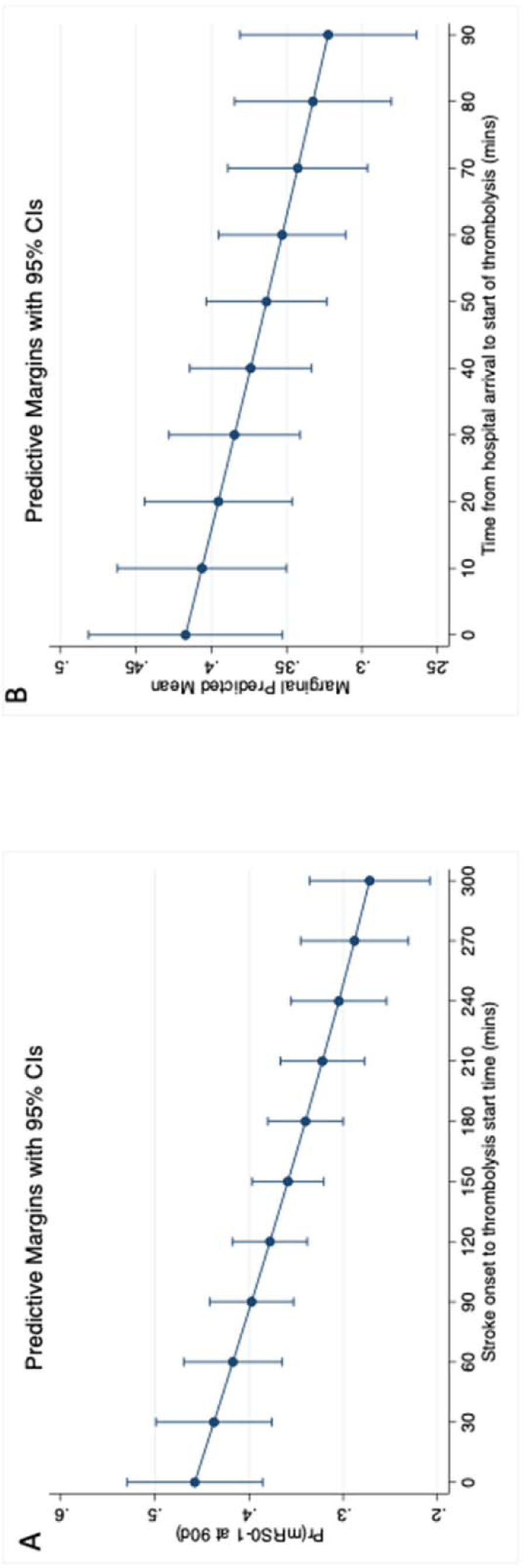
Relationship between onset to needle time and mRS 0-1 at 90-120 days using mixed effects logistic regression model adjusted for age, sex, stroke severity as measured using the National Institute of Health Stroke Scale (NIHSS) and intracranial occlusion site as fixed effects variables and “enrolling site” as the random effects variable (error bars indicate 95% CI) (A). It shows that for every 30-minute reduction in onset to needle time two more out of a 100 people achieve an excellent outcome. Relation between door to needle time and mRS 0-1 at 90-120 days using mixed effects logistic regression model adjusted for age, sex, stroke severity and occlusion site with site as random effects (error bars indicate 95% CI) (B). It shows that for every 60-minute reduction in door to needle time one more out of a 100 people achieve an excellent outcome.

